# Multimodal Integration of Alzheimer’s Plasma Biomarkers, MRI, and Genetic Risk for Individual Prediction of Cerebral Amyloid Burden

**DOI:** 10.1101/2025.05.07.25326855

**Authors:** Yichen Wang, Lairai H.J. Chen, Yuxin Cheng, Yuyan Cheng, Shiyun Zhao, Yidong Jiang, Tianyu Bai, Yanxi Huo, Kexin Wang, Mingkai Zhang, Weijie Huang, Guozheng Feng, Ying Han, Ni Shu

## Abstract

Alzheimer’s disease (AD), the most prevalent neurodegenerative disorder, is marked by the accumulation of amyloid-β (Aβ) plaques. Although cerebral Aβ positron emission tomography (Aβ-PET) remains the gold standard for assessing cerebral Aβ burden, its clinical utility is hindered by cost, radiation exposure, and limited availability. Plasma biomarkers serve as promising non-invasive predictors of cerebral Aβ burden, but reliance on a single marker often leads to suboptimal predictive performance. To address this, we proposed a multimodal machine learning strategy that integrates readily accessible and non-invasive features—such as plasma biomarkers, structural magnetic resonance imaging (sMRI)-derived atrophy measures, diffusion tensor imaging (DTI)-based structural connectomes (SCs), and genetic risk profiles—to predict cerebral Aβ burden and evaluate the relative contribution of each modality to predictive performance. Specifically, a random forest regressor was trained using data from the Alzheimer’s Disease Neuroimaging Initiative (ADNI; n = 150) and evaluated with leave-one-out cross-validation. Our results showed that integrating multimodal features improves the predictive power on cerebral amyloid burden: while the baseline model using plasma and clinical variables alone achieved an R² of 0.52, adding neuroimaging and apolipoprotein E (APOE) genotype features improved performance (R² = 0.617), and replacing APOE with polygenic risk scores (PRS) further enhanced accuracy (R² = 0.637). The predictive value of multimodal integration was also replicated in an independent cohort (SILCODE; n = 101). Moreover, a multiclass classifier trained with the same multimodal features achieved high accuracy in distinguishing clinical stages of Aβ burden—normal controls (NC), mild cognitive impairment (MCI), and Alzheimer’s disease (AD)—with area under the curve (AUC) values of 0.86, 0.77, and 0.93, respectively. These findings highlight the value of combining plasma, imaging, and genetic data to non-invasively estimate cerebral Aβ burden, offering a potential alternative to PET imaging for early AD risk assessment.

## 1 Introduction

Alzheimer’s disease (AD) is the most prevalent neurodegenerative disorder, characterized by the accumulation of amyloid-β (Aβ) plaques and neurofibrillary tangles, both of which are closely linked to cognitive decline^1,2^. Aβ positron emission tomography (Aβ-PET) remains the gold standard for in vivo quantification of cerebral Aβ burden, providing region-specific estimates of amyloid deposition^3^. However, its high cost, invasive nature, and limited accessibility hinder its applicability for large-scale or routine individual screening.

Plasma biomarkers—including phosphorylated tau (reflecting tau hyperphosphorylation), β-amyloid 42/40 ratio (Aβ_42_/Aβ_40_; indexing amyloid burden), neurofilament light chain (NfL; indicating axonal injury), and glial fibrillary acidic protein (GFAP; reflecting astrocytic activation)—have shown promise as accessible, non-invasive indicators for estimating cerebral Aβ burden^4^. However, their standalone predictive performance remains limited, particularly at the individual level, due to biological heterogeneity and the indirect nature of peripheral measurements. Structural MRI (sMRI) and diffusion tensor imaging (DTI) provide non-invasive insights into brain changes associated with Alzheimer’s disease^5^. sMRI characterizes regional atrophy, whereas DTI-derived structural connectomes (SCs), capturing inter-regional white matter connectivity^6,7^.

Genetic factors also modulate the predictive capacity of plasma biomarkers. The apolipoprotein E (APOE) ε4 allele, the most established genetic risk factor for AD^8^, is easily genotyped and widely used in clinical settings. Polygenic risk scores (PRS), although theoretically more comprehensive by aggregating genome-wide variants, are often limited by their cost and complexity^9^. Both may provide complementary information beyond plasma biomarkers^10,11^. However, given the higher cost of PRS assessment, it is necessary to consider the feasibility of using APOE genotype as a cost-effective proxy.

Together, plasma, neuroimaging, and genetic biomarkers capture distinct yet complementary dimensions of Alzheimer’s disease pathophysiology^12^. However, integrating these heterogeneous data sources remains challenging due to their different dimensionality and complex interdependencies. Among machine learning approaches, random forest (RF) algorithms are particularly well-suited for modeling multimodal biomedical data^13^. RFs can effectively accommodate nonlinear relationships, and provide interpretable feature importance metrics, making them ideal for individualized prediction of cerebral Aβ burden.

Previous studies have explored Aβ burden prediction using selected data modalities, but few have integrated multiple complementary biological domains, such as plasma, imaging, and genetics. For example, Ramanan et al. demonstrated that adding genetic risk scores to plasma biomarkers improved diagnostic accuracy for brain amyloidosis^10^. Santos et al validated the utility of plasma biomarkers for predicting dementia conversion in a Brazilian cohort^14^. Other studies have utilized machine learning on imaging and cognitive features to identify incipient dementia cases^15^, while some reviews have summarized available plasma assays and highlighted their promise for clinical translation^16^. However, these efforts typically focus on one or two modalities. A truly integrative multimodal framework—combining plasma, neuroimaging, and genetic data—is still lacking. Furthermore, no prior study has systematically evaluated the relative contribution of each modality within a unified predictive framework, leaving it unclear which data sources offer the most substantial incremental value.

To address these gaps, we proposed a multimodal machine learning framework based on ML to predict continuous cerebral Aβ burden. Specifically:

(1) We evaluated the model using data from the ADNI, systematically examining the added predictive value of structural MRI, DTI-derived structural connectomes, and genetic features beyond plasma biomarkers; (2) We compared the predictive performance of APOE genotype and PRS within this framework to evaluate their respective utility. (3) We validated the model in an independent Chinese cohort (SILCODE) to assess cross-cohort generalizability

## 2 Methods

### 2.1 Participants and Study Design

This study conducted a secondary analysis using publicly available data from the ADNI, a longitudinal project aimed at identifying early biomarkers of AD through multimodal data collection. ADNI participants were included if they were aged 55–90 years, generally in good health, fluent in English or Spanish, and had a Geriatric Depression Scale score below 6. Clinical diagnoses—cognitively unimpaired (CU), mild cognitive impairment (MCI), or AD—were determined according to standard ADNI criteria based on subjective memory complaints, objective neuropsychological assessments, and clinician-rated global functioning. To ensure multimodal data availability, participants with sMRI, DTI, genetic data, plasma biomarkers, and Aβ-PET imaging acquired within a one-year interval were selected from the ADNI-GO, ADNI-2, and ADNI-3 phases.

To assess the generalizability, we performed external validation using the SILCODE cohort—a prospective multicenter study of AD and cognitive impairment in the mainland Chinese population. SILCODE participants were included if they were aged 45–90 years, right-handed, native Mandarin speakers, and had complete multimodal data for sMRI, DTI, plasma biomarkers, APOE genotyping, and Aβ-PET imaging within a one-year window. Demographic characteristics of both cohorts, stratified by diagnostic group, are summarized in **Table 1**.

**Table 1.** Demographic, clinical, imaging, and biomarker characteristics of participants in the ADNI and SILCODE cohorts, stratified by diagnostic group: cognitively unimpaired (CU), mild cognitive impairment (MCI), and Alzheimer’s disease (AD). Values are reported as mean ± standard deviation or N (%). PET_CL: Centiloid values from amyloid PET; SUVR: standardised uptake value ratio from amyloid PET; Pla_tau217 and Pla_tau181: plasma phosphorylated tau at threonine 217 and 181, respectively; Pla_Aβ42/Aβ40: plasma amyloid-β 42/40 ratio; Pla_NfLQ: plasma neurofilament light chain; Pla_GFAP: plasma glial fibrillary acidic protein; APOE_e4: apolipoprotein E ε4 dosage (range: 0–2); PRS_1e-6: polygenic risk score based on SNPs with p < 1×10□□; Education: years of formal education; MMSE: Mini-Mental State Examination score. “\” indicates variables not available in the corresponding cohort or subgroup.

|  | ADNI |  |  | SILCODE |  |  |
| --- | --- | --- | --- | --- | --- | --- |
| Diagnose | CU | MCI | AD | CU | MCI | AD |
| N | 97 | 43 | 10 | 57 | 24 | 20 |
| Age | 71.36±6.02 | 74.84±8.62 | 72.27±6.57 | 68.16±5.71 | 70.46±6.31 | 71.40±10.98 |
| Female | 36(37%) | 26(60%) | 7(70%) | 40(61%) | 14(42%) | 12(60%) |
| **Pet_CL** | 21.81±31.15 | 44.53±54.78 | 102.10±46.53 | \ | \ | \ |
| **SUVr** | \ | \ | \ | 1.03±0.11 | 1.09±0.16 | 1.3±0.17 |
| **Pla_tau217** | 0.21±0.27 | 0.34±0.34 | 0.73±0.38 | \ | \ | \ |
| **Pla_tau181** | \ | \ | \ | 2.04±0.94 | 2.82±1.04 | 3.79±1.17 |
| **Pla_Aβ42/Aβ40** | 0.09±0.01 | 0.13±0.2 | 0.08±0.01 | 0.06±0.02 | 0.06±0.01 | 0.05±0.02 |
| **Pla_NfLQ** | 19.56±8.7 | 23.24±12.84 | 25.58±8.55 | 17.64±7.1 | 22.09±7.71 | 31.62±22.82 |
| **Pla_GFAP** | 159.96±78.9 | 172.83±107.16 | 244.32±114.55 | 102.32±44.49 | 118.08±70.02 | 164.09±114.95 |
| **APOE_e4** | 0.41±0.63 | 0.65±0.65 | 1±0.67 | 0.23±0.46 | 0.5±0.59 | 0.55±0.51 |
| **PRS_1e-6** | -0.16±0.94 | 0.12±0.89 | 0.25±0.62 | \ | \ | \ |
| **Education** | 16.93±2.28 | 15.63±2.55 | 15.6±1.6 | 14.51±3.24 | 13.29±2.8 | 11.75±4.24 |
| **MMSE** | 29.24±1.17 | 26.63±2.9 | 23.4±2.17 | 28.42±1.7 | 24.54±2.38 | 18.35±6.39 |

### 2.2 MRI Data Acquisition

sMRI and DTI data were collected from both the ADNI and SILCODE cohorts. In ADNI, MRI scans were acquired using 3.0 Tesla (3T) scanners from Philips, Siemens, or GE Healthcare across the ADNI-GO, ADNI-2, and ADNI-3 phases. For sMRI, a standardized protocol was used, with typical parameters including a repetition time (TR) of 6.7 ms, echo time (TE) of 3.1 ms, flip angle of 9°, field of view (FOV) of 256 × 256 × 170 mm³, slice thickness of 1.2 mm, and voxel size of 1.0–1.11 mm^³^, covering 170 sagittal slices. DTI scans in ADNI-GO and ADNI-2 were acquired using GE 3T scanners with TR = 9000 ms, TE = 60–70 ms, and voxel size = 1.37 × 1.37 × 2.7 mm³. Each scan included 30 diffusion-weighted directions (b = 1000 s/mm^²^) and 5 non-diffusion-weighted (b = 0) images. For ADNI-3, an updated protocol was used with a resolution of 2 × 2 × 2 mm³, TR = 7200 ms, TE = 56 ms, 35 axial slices, and a total scan time of approximately 7.5 minutes.

In the SILCODE cohort, imaging was performed using a 3T integrated PET/MR scanner (SIGNA PET/MR, GE Healthcare) at Xuanwu Hospital, Capital Medical University. sMRI was acquired using a gradient-recalled echo sequence with TR = 2300 ms, inversion time (TI) = 900 ms, TE = 2.26 ms, flip angle = 8°, FOV = 256 × 256 mm^²^, slice thickness = 1 mm, and isotropic voxel size of 1 mm^³^. DTI data were collected using a spin-echo echo-planar imaging (EPI) sequence with TR = 4500 ms, TE = 65 ms, voxel size = 2 × 2 × 2 mm^³^, and 64 slices. Diffusion gradients were applied in 128 directions, including 64 at b = 1000 s/mm² and 64 at b = 2000 s/mm², along with one non-diffusion-weighted (b = 0) image.

### 2.3 MRI Data Processing

All imaging data were preprocessed using standardized protocols adapted from the UK Biobank pipeline^17^, implemented using *FSL*^18^, *FreeSurfer*^19^, and *MRtrix3*^20^. Raw DICOM images were converted to NIfTI format using *dcm2niix*^21^. For diffusion imaging, b-value distributions were inspected prior to preprocessing. sMRI preprocessing for both cohorts included skull stripping and bias field correction using FSL, followed by cortical reconstruction and volumetric segmentation using FreeSurfer v6.0. This yielded native-space parcellations based on the Desikan–Killiany atlas. These parcellations were further used to extract mean SUVR values within regions of interest (ROIs) for each participant. DTI data were corrected for head motion, eddy currents, and EPI distortions using FSL’s eddy^22^.

SCs were constructed using a unified tractography pipeline. Fiber orientation distributions were estimated using constrained spherical deconvolution, followed by probabilistic tractography^23^. Tractography was seeded from the gray–white matter interface using the *5ttgmwmi* algorithm^24^. Brain network nodes were defined using a hybrid atlas comprising 100 cortical regions from the Schaefer atlas^25^ and 16 subcortical regions from the Melbourne Subcortex Atlas (MSA) ^26^. The resulting SC matrices were used for feature extraction and predictive modeling. To further characterize topological changes at the network level, we grouped the 116 regions into 8 functional modules based on Yeo’s 7-network parcellation scheme and the MSA-defined subcortical network. The Yeo-7 system defines seven large-scale cortical functional networks: visual, somatomotor, default mode, dorsal attention, salience/ventral attention, executive control and limbic networks. The eighth module, subcortical, includes 16 subcortical nuclei defined by the MSA.

### 2.4 Quantification and Harmonization of Amyloid PET Measures

In the ADNI cohort, Aβ burden was quantified using Centiloid (CL) values ^27,28^, processed uniformly by the University of California, Berkeley, and publicly released alongside the official dataset. CL values were derived from cortex-wide *SUMMARY_SUVR* measures, normalized to the whole cerebellum, and converted using tracer-specific formulas:

For [¹□F]-florbetapir (FBP):

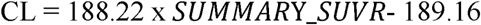

For [¹□F]-florbetaben (FBB):

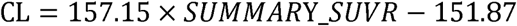

*SUMMARY_SUVR* values were based on sMRI-guided segmentation of four cortical regions (frontal, cingulate, parietal, and temporal), with normalization to the whole cerebellum; regional definitions are detailed in the Supplementary Materials (Table S1). In ADNI cohort, we utilized the standardized CL values provided by ADNI rather than recalculating SUVRs. This decision was based on two considerations: (i) the CL framework enables cross-tracer and cross-scanner harmonization, improving comparability and robustness; and (ii) the official CL pipeline has been extensively validated and widely adopted, ensuring high reproducibility.

In the SILCODE cohort, Aβ PET imaging was conducted using the radiotracer [^18^F]-florbetapir (AV-45). Participants received an intravenous injection of 7 to 10 mCi of [^18^F]-florbetapir, followed by a rest period of approximately 40 min, after which a 20-minute static PET scan was performed. PET data acquisition employed a time-of-flight ordered subset expectation maximization (TOF-OSEM) algorithm^29,30^ with 8 iterations, a field of view of 350 × 350 mm2, subset matrices of 32 at 192 × 192, and a full width at half maximum (FWHM) of 3 mm. Aβ burden was quantified using *SUMMARY_SUVR* values, calculated with the same cortical and reference regions as in the ADNI processing pipeline.

### 2.5 Genetic Risk Assessment

In ADNI, genotyping was performed using either the Illumina HumanOmniExpress or Infinium Global Screening Array v2. Each dataset underwent separate preprocessing, imputation, and quality control (QC) steps to ensure batch consistency. Preprocessing involved recoding chromosome labels, removing non-autosomal variants and duplicate SNPs, and correcting strand orientations using PLINK^31^ and Bcftools^32^. Phasing used the 1000 Genomes Phase 3 panel^33^, with imputation via Impute5^34^. Variants with imputation INFO scores < 0.8 were excluded. After merging batches, unified QC removed SNPs with minor allele frequency < 0.01, genotype missingness > 1%, or significant deviation from Hardy–Weinberg equilibrium (p < 1×10^-6^). Individuals were excluded for high genotype missingness, excessive heterozygosity (F-statistic ±3 SD), cryptic relatedness (identity-by-descent > 0.125), or discrepancies between genetically inferred and reported sex. PRS were computed using clumping and thresholding (C+T) based on AD GWAS summary statistics^35^, testing multiple p-value inclusion thresholds (p < 10^-6^) combined with LD-based clumping (window size=250 kb, r²<0.01). Resulting scores were standardized (Z-scores) for analysis. To control population stratification, the top five genetic principal components derived via PCA were included as covariates. In SILCODE, genome-wide genotyping was unavailable. Only APOE genotype data obtained through standardized laboratory protocols were included as categorical covariates (ε2, ε3, ε4) in analyses.

### 2.6 Multimodal Feature Integration and Predictive Modeling

To build an accurate prediction model and assess the added predictive value of sMRI, SC, and genetic features beyond plasma biomarkers, we implemented a modular machine learning framework enabling multimodal integration and systematic comparison across feature combinations^10,11,36^. Taking plasma features as the baseline, we defined and compared five feature combinations: Clinical + Plasma, Clinical + Plasma + APOE, Clinical + Plasma + SC, Clinical + Plasma + MRI, and Clinical + Plasma + MRI + SC + APOE. Two regression algorithms, linear regression and random forest (RF) regression, were used for model benchmarking (Fig. 1).

**Figure 1.**
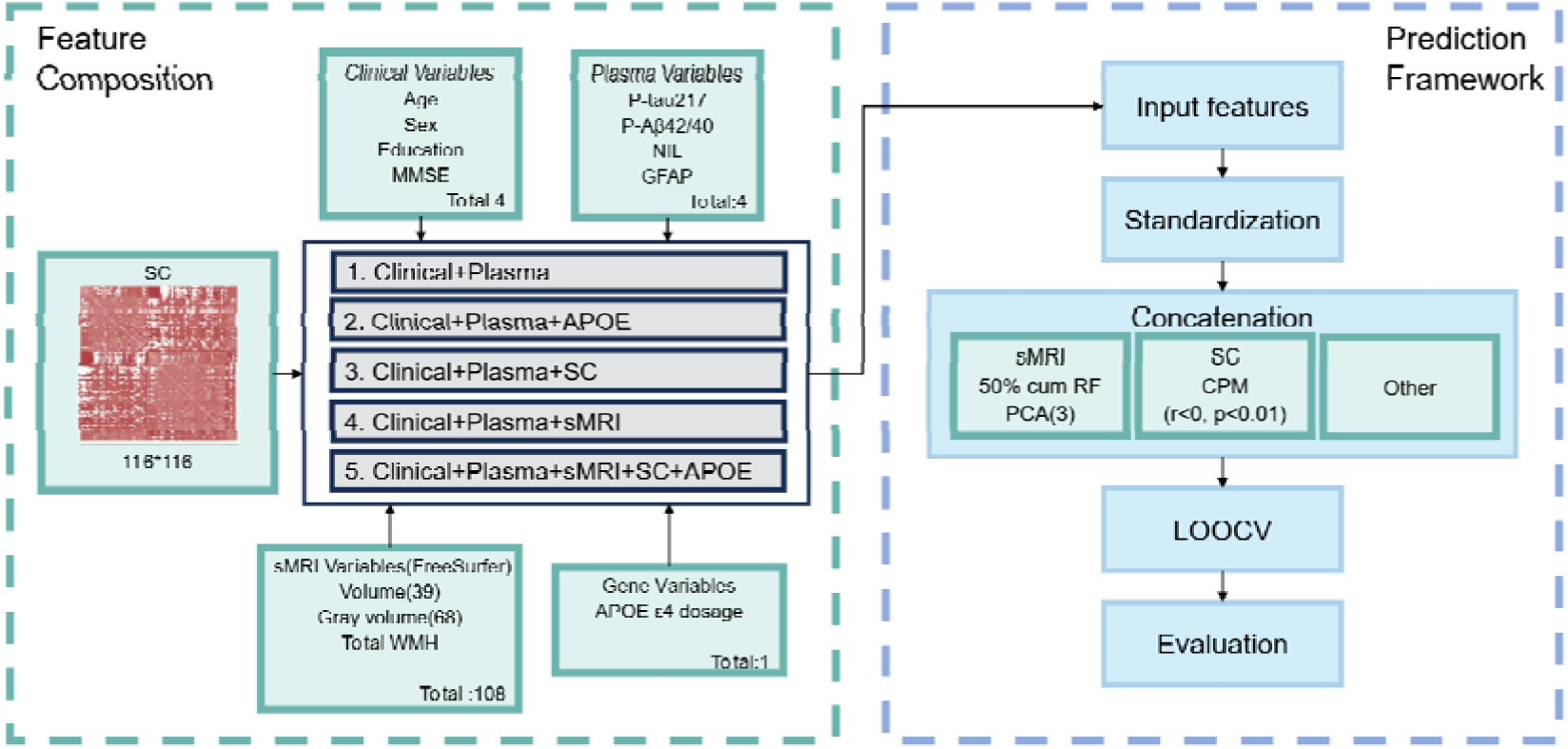
Multimodal machine learning framework for predicting cerebral amyloid-β burden. An integrative pipeline was constructed to combine clinical, plasma, genetic, sMRI, and SC features. Five feature combinations were tested under a LOOCV framework. sMRI features and SC were dimensionally reduced or selected based on established criteria. Models were trained using both linear and random forest regressors. Abbreviations: Aβ, amyloid-β; APOE, apolipoprotein E4 dosage; CPM, connectome-based predictive modeling; DTI, diffusion tensor imaging; LOOCV, leave-one-out cross-validation; PCA, principal component analysis; PRS, polygenic risk score; SC, structural connectome; sMRI, structural MRI; MMSE, Mini-Mental State Examination.

A total of 108 sMRI features were included, encompassing cortical volumes, surface areas, and white matter hyperintensities (WMHs), all normalized by intracranial volume (ICV) to control for inter-individual differences in brain size^37^. Within each training set, features were ranked by importance using RF regression, with the top 50% retained and further reduced to three orthogonal components using PCA. The SC features were extracted using the CPM (connectome-based predictive modeling) approach^38,39^. For each training fold, edges negatively correlated with CL values (Pearson’s r < 0, p < 0.01) were identified.

The average strength of these negatively correlated edges was computed as a summary feature per subject.

Model training and validation were performed using leave-one-out cross-validation (LOOCV)^40^. All preprocessing, feature selection, and dimensionality reduction steps were strictly confined to the training data in each fold. The resulting model was then applied to the held-out subject to obtain unbiased performance estimates. The ML framework modelling was conducted separately within ADNI and SILCODE.

To further evaluate model performance in classifying clinical Aβ burden status, we trained multi-class Random Forest classifiers using sing clinical diagnosis labels (NC, MCI, AD) as outcomes. Classification was conducted in ADNI cohort under the same LOOCV framework, and model performance was assessed using area under the receiver operating characteristic curve (AUC) for each class, as well as macro-average AUC to evaluate overall discriminative ability. The same feature extraction and selection pipeline was applied as in the regression setting, ensuring consistent input modalities across tasks.

### 2.7 Interpretability Analysis

To further elucidate the contributions of individual features to model predictions and enhance interpretability, we employed SHapley Additive exPlanations (SHAP)^41^ to interpret the predictive power of each feature. SHAP quantifies the marginal contribution of each input feature to a given prediction while accounting for feature interactions by constructing a local linear model to approximate and interpret complex nonlinear models *f(x)*, formulated as:

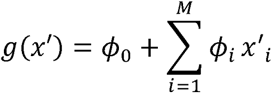

where *x’* represents a simplified binany version of the input features (indicating feature presence or absence), *ϕ_O_* denotes the expected output when all features are absent, and *ϕ*_i_ represents the Shapley value of feature *i*, reflecting its marginal contribution to the prediction.

### 2.8 Sensitivity Analysis

To evaluate the robustness of the proposed framework, we conducted sensitivity analyses across three methodological dimensions: (1) PRS construction, and (2) SC parcellation templates.

First, PRSs were reconstructed using clumping and thresholding with a range of p-value cutoffs (p□<□10^-5^, 10□^6^ 10^-7^, 10^-8^, 10^-9^, and 10^-10^). Second, to evaluate the effect of brain parcellation schemes, we derived SC features using an alternative atlas combining aparc and MSA-16 (total 84 regions), replacing the default Schaefer-100 + MSA-16 template (total 116 regions).

## 3 Results

### 3.1 Associations between features and cerebral amyloid-β burden

In the ADNI cohort, among plasma biomarkers, p-tau_217_ showed the strongest positive association with cerebral Aβ burden (β = 0.591, p = 1.70 × 10^-15^), followed by GFAP (β = 0.454, p = 5.58 × 10□□) and NfL (β = 0.217, p = 7.73 × 10□³). In contrast, the Aβ_42_/Aβ_40_ ratio was strongly and negatively associated with Aβ burden (β = –4.579, p = 5.32 × 10□¹²). Cognitive performance, as measured by MMSE, exhibited a modest negative association with Aβ burden (β = –0.172, p = 2.27 × 10□□), suggesting that amyloid deposition contributes only partially to cognitive decline. Compared to plasma markers, genetic features showed weaker associations: the PRS was positively correlated with Aβ burden (β = 0.262, p = 1.19 × 10□³), while APOE ε4 dosage revealed a clear dose-dependent increase in amyloid levels (Fig. 2a–b). Structural MRI analyses revealed multiple brain regions associated with Aβ burden. The eight regions with the strongest correlations, as shown in Figure 2c, were mainly located in the medial temporal lobe (e.g., amygdala, hippocampus), parietal cortex (e.g., precuneus, inferior parietal lobule), and subcortical areas. These spatial patterns are consistent with regions known to be vulnerable in the early stages of Alzheimer’s disease, reinforcing the link between structural atrophy and amyloid deposition.

**Figure 2.**
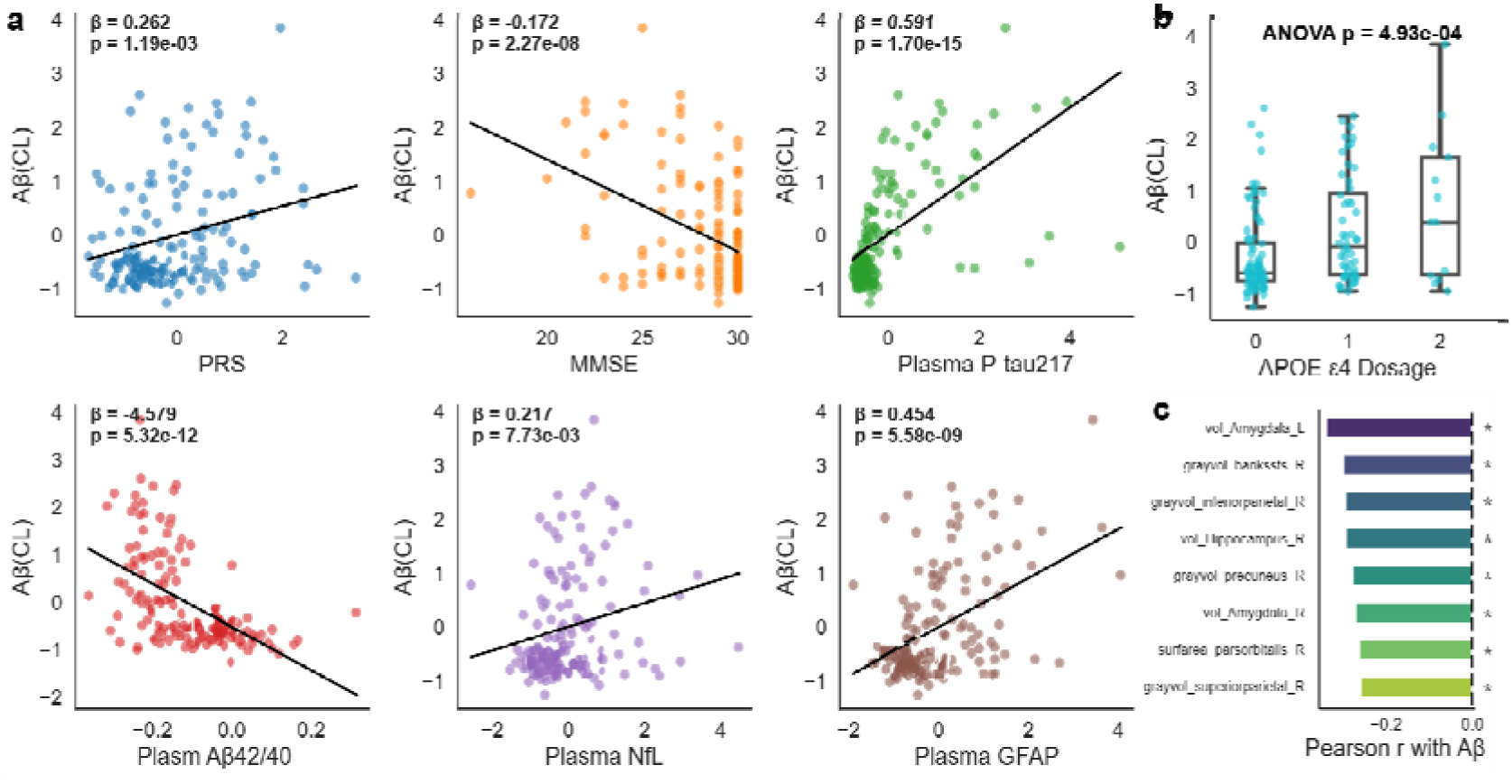
Associations between plasma biomarkers, cognitive function, genetic risk, and brain structural features with standardized Aβ burden. (a) Scatter plots showing the linear associations between Z-scored CL values and PRS, MMSE, p-tau_217_, Aβ_42_/Aβ_40_ ratio, NfL, and GFAP. Standardized regression coefficients (β) and corresponding p-values are annotated in each panel. (b) Boxplots illustrating the distribution of Z-scored CL values stratified by APOE ε4 allele dosage (0, 1, or 2 copies). (c) The top eight ICV-normalized sMRI features (volume or surface area) most strongly correlated with Aβ burden, ranked by Pearson correlation coefficients. Asterisks (*) indicate significant associations (p < 0.05, uncorrected). Abbreviations: Aβ, amyloid-β; APOE, apolipoprotein E; CL, Centiloid; GFAP, glial fibrillary acidic protein; MMSE, Mini-Mental state Examination; NfL, neurofilament light chain; PRS, polygenic risk score; ICV, Intracranial Volume; ANOVA, analysis of variance.

### 3.2 Multimodal feature integration enhances Aβ burden prediction performance

We systematically evaluated the predictive performance of different feature combinations under a LOOCV framework using data from the ADNI cohort. Linear regression showed limited ability to model cerebral Aβ burden across all feature sets (R^2^ = 0.38 for the Clinical + Plasma + sMRI + SC + APOE model), indicating its inability to capture the complex, nonlinear interactions inherent to multimodal data. In contrast, the RF model consistently outperformed linear regression, with performance improving as more modalities were integrated (Fig. 3a–b). Notably, the full model incorporating clinical features, plasma biomarkers, sMRI, SC, and APOE genotype achieved the best predictive accuracy (R² = 0.617) and lowest mean squared error (MSE = 770.1) (Fig. 3c), highlighting the synergistic value of combining diverse biological information.

**Figure 3.**
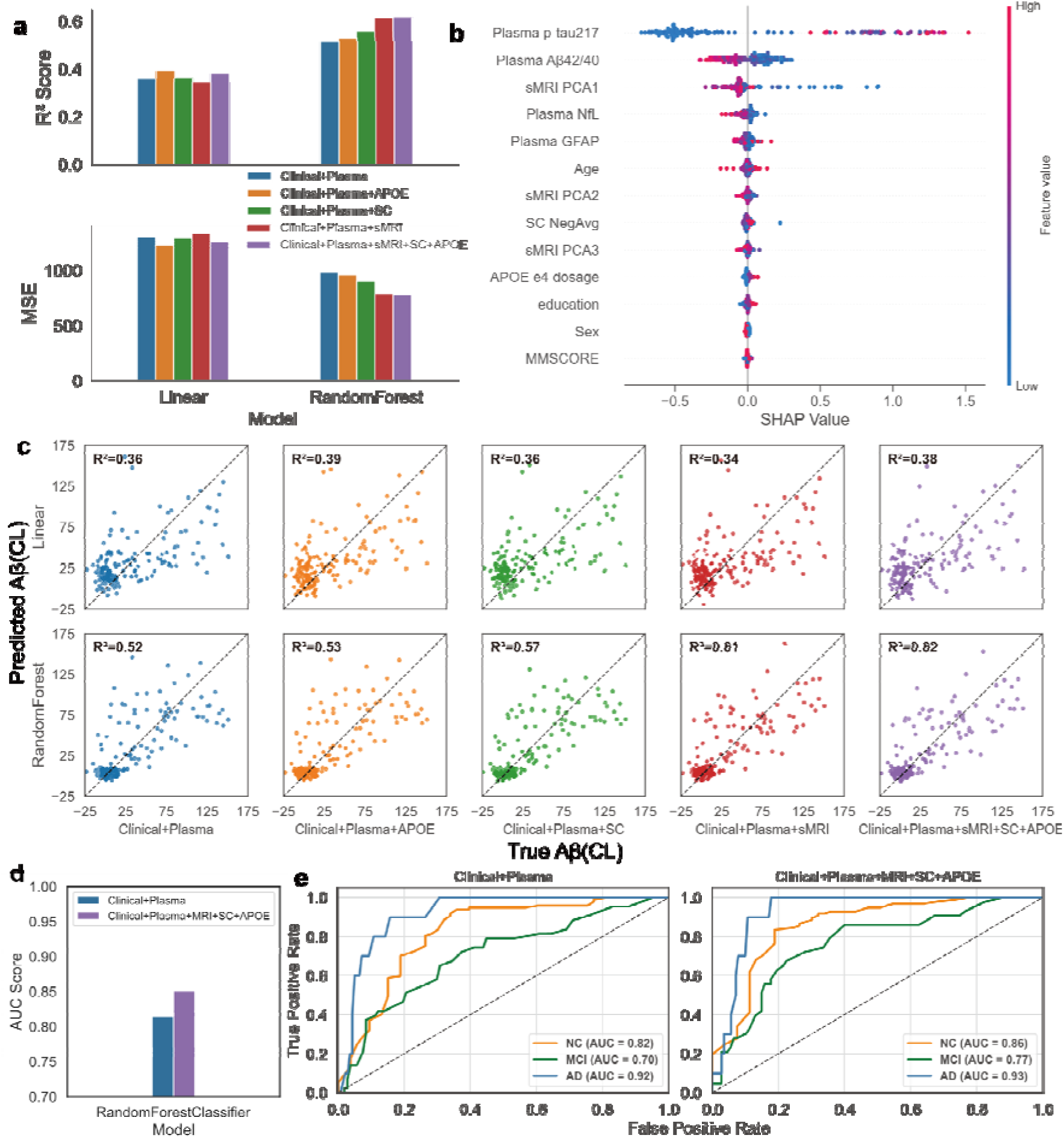
Model performance and SHAP-based feature importance for Aβ burden prediction in the ADNI cohort. (a) R² and mean squared error (MSE) for linear regression (LR) and random forest (RF) models across different feature combinations under leave-one-out cross-validation (LOOCV). (b) SHAP summary plot showing the top features in the best-performing RF model (Clinical + Plasma + sMRI + SC + APOE), ranked by mean absolute SHAP value. Each point represents a subject, with colour indicating feature value (red = high, blue = low). (c) Scatter plots of observed versus predicted Aβ burden (Centiloid values) for LR and RF models across all feature combinations. R² values are shown for each model. (d) Macro-average area under the receiver operating characteristic curve (AUC) for multi-class classification (NC, MCI, AD) across different feature combinations using RF classifier. (e) Class-specific AUC values for NC, MCI, and AD groups based on RF models trained with various combinations of features. Abbreviations: Aβ, amyloid-β; APOE, apolipoprotein E; GFAP, glial fibrillary acidic protein; MMSE, Mini-Mental State Examination; NfL, neurofilament light chain; PCA, principal component analysis; SC, structural connectivity; SHAP, SHapley Additive exPlanations; RF, random forest; LR, linear regression; MSE, mean squared error; ROC, receiver operating characteristic; AUC, area under the curve; NC, cognitively normal; MCI, mild cognitive impairment; AD, Alzheimer’s disease; Clinical, demographic and cognitive measures.

Specifically, the RF model using only clinical and plasma features achieved an R^2^ of 0.515. Incremental gains were observed with the addition of APOE genotype (R^2^ = 0.527), SC features (R^2^ = 0.556), and sMRI measures (R^2^ = 0.615), with maximal performance attained when all modalities were combined (R^2^ = 0.617). Detailed R² and MSE metrics for all feature sets are provided in Supplementary Table S2.

To interpret the model, we applied SHAP analysis to the best-performing RF model. The most influential predictors were plasma biomarkers, particularly p-tau_217_ and the Aβ_42_/Aβ _40_ ratio, followed by principal components from sMRI, SC-derived connectivity metrics and genetic feature. These results suggest that while plasma biomarkers offer strong individual predictive power, imaging and genetic feature provide valuable complementary information.

To assess classification performance across clinical stages, we further trained a multi-class RF classifier using clinical diagnosis labels (NC, MCI, AD). Compared to the baseline model using clinical and plasma features (AUC = 0.82 for NC, 0.70 for MCI, and 0.92 for AD), the full multimodal model improved classification performance across all subgroups (AUC = 0.86 for NC, 0.77 for MCI, and 0.93 for AD) (Fig. 3e). Macro-average AUC also improved (Fig. 3d), reinforcing that multimodal integration enhances both continuous and categorical prediction of cerebral amyloid pathology.

### 3.3 Comparing the predictability of APOE and PRS

To evaluate the additive predictive value of genetic information, we compared the performance of APOE genotype and PRS in predicting cerebral Aβ burden within the ADNI cohort (Fig. 4). In the baseline model comprising clinical and plasma biomarkers, the inclusion of APOE ε4 status led to a modest improvement in prediction accuracy (ΔR² = 0.012). In contrast, substituting APOE with PRS yielded a more substantial gain (ΔR² = 0.056), suggesting a broader capture of genetic risk beyond the APOE locus. A detailed comparison is provided in Supplementary Materials (Table S3).

**Figure 4.**
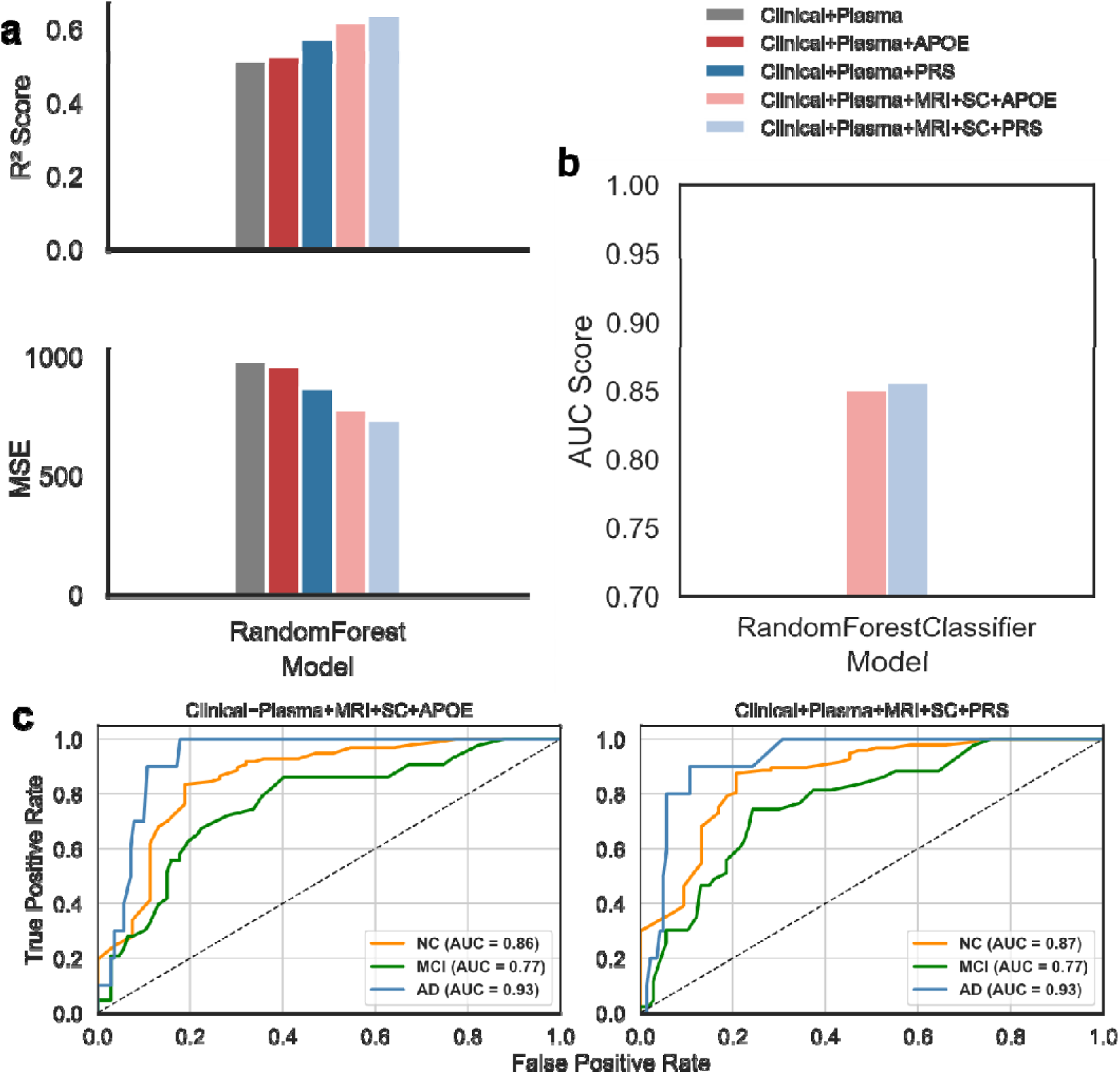
Model performance with APOE or PRS for predicting Aβ burden in the ADNI cohort. Bar plots display R² (top) and MSE (bottom) for models using different combinations of clinical, plasma, genetic, and imaging features. (b) AUC scores from multi-class Random Forest classifiers trained using CL-based Aβ labels (NC, MCI, AD) with either APOE or PRS added to the baseline Clinical + Plasma + sMRI + SC model. (c) ROC curves for each class (NC, MCI, AD) derived from models using APOE (left) or PRS (right) in the full feature combination.

This advantage of PRS persisted when more complex models incorporating structural MRI and structural connectivity features were used. Although the performance gap between APOE and PRS narrowed in the full multimodal model (R²_APOE_ = 0.617 vs. R²_PRS_ = 0.637), PRS consistently demonstrated higher predictive accuracy across all combinations.

This trend was also reflected in classification performance. As shown in Fig. 4b–c, both APOE- and PRS-based models achieved similarly high AUCs in distinguishing between NC, MCI, and AD groups based on CL-derived Aβ burden. The PRS-based model achieved AUCs of 0.87 (NC), 0.77 (MCI), and 0.93 (AD), closely matching the APOE-based model (0.86, 0.77, and 0.93, respectively). These results indicate that while PRS provides superior continuous-level prediction of Aβ burden, its classification performance is comparable to APOE when stratifying disease stages.

### 3.4 Distributed Network Disruption Associated with Aβ Burden

To identify SC consistently weakened by Aβ burden, we applied CPM under a LOOCV framework. In each fold, connections negatively associated with CL values (p□<□0.01) were selected, and those recurring across all folds were retained, yielding 82 stable disconnections (Fig. 5). To identify SC consistently weakened by Aβ burden, we applied CPM under a LOOCV framework. In each fold, connections negatively associated with CL values (p□<□0.01) were selected, and those recurring across all folds were retained, yielding 82 stable disconnections (Fig. 5).

**Figure 5.**
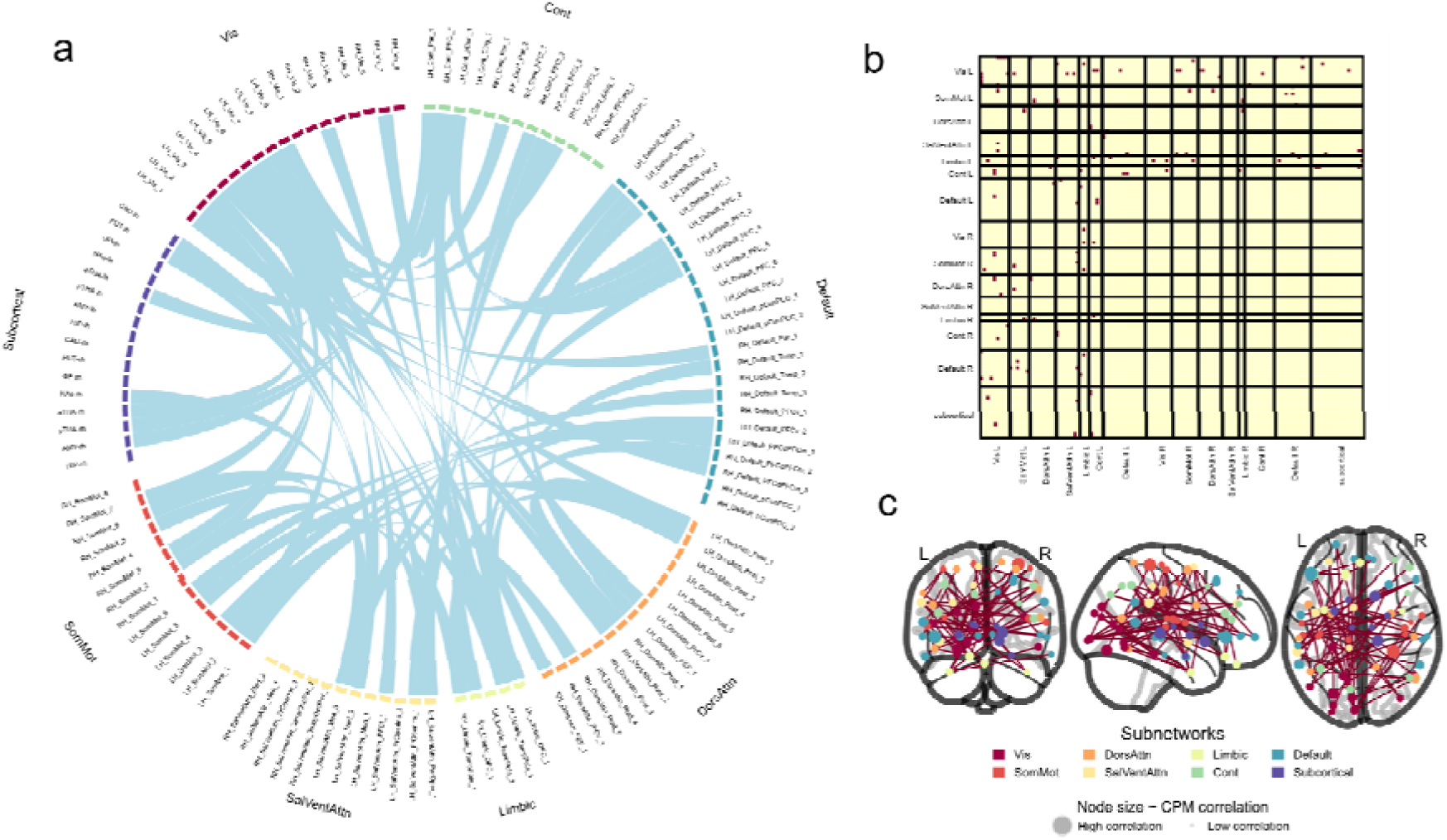
Structural connections with common associations with CL value across all folds in ADNI. (a) Chord diagram illustrating the intersection of significantly positive connections identified by CPM across all training sets under the LOOCV framework. Nodes are grouped by functional subnetworks. (b) Inter-subnetwork connectivity matrix displaying the distribution of stable connections among major functional modules. Red dots indicate the presence of consistently selected edges; rows and columns represent different subnetworks. (c) Cortical surface projection of stable structural connections. Node size indicates CPM correlation strength, and both nodes and edges are colour-coded by functional subnetwork. Abbreviations: CPM, connectome-based predictive modeling; LOOCV, leave-one-out cross-validation; Vis, visual network; SomMot, somatomotor network; DorsAttn, dorsal attention network; SalVentAttn, salience/ventral attention network; Limbic, limbic network; Default, default mode network; Cont, control network; Subcortical, subcortical structures.

Among all functional subnetworks, the visual network exhibited the highest number of disconnections, both within itself (n = 10) and in its connections with other networks—particularly the control (n = 5), somatomotor (n = 4), and default mode (n = 4) networks. This dominant pattern of disconnection suggests early vulnerability of perceptual and attentional integration systems during amyloid accumulation.

In addition to the visual network, spatial mapping of affected regions revealed consistent involvement of the medial prefrontal cortex and lateral parietal cortex—key hubs of the default mode and frontoparietal control networks. These findings indicate that Aβ-related disruptions extend beyond primary sensory systems and impact higher-order cognitive networks implicated in early-stage AD.

Collectively, the observed disconnection patterns support a network-level framework for understanding AD, wherein early amyloid deposition perturbs distributed integrative systems. Structural connectome features derived from robust CPM analysis may thus serve as sensitive and interpretable biomarkers for early disease characterization and risk stratification.

### 3.5 External validation in the SILCODE cohort

To assess generalizability, we performed external validation using the SILCODE cohort, employing five feature combinations with RF models to predict Aβ PET SUVR (Fig. 6). Relative to the baseline model (Clinical+Plasma; R²=0.232), incremental performance improvements were observed upon adding APOE genotype (ΔR²=+0.009), SC features (ΔR²=+0.027), and sMRI features (ΔR²=+0.032). The full model (Clinical+Plasma+MRI+SC+APOE) achieved the highest R² of 0.267, a detailed comparison of R² and MSE across models is provided in Supplementary Materials (Table S4). Although overall predictive accuracy was lower than in ADNI, the incremental benefit pattern remained consistent, highlighting multimodal robustness. Lower performance in SILCODE primarily reflected plasma biomarker differences: ADNI utilized plasma p-tau_217_, known for superior amyloid sensitivity, whereas SILCODE employed the less sensitive but widely used p-tau_181_. Prior studies have confirmed that p-tau_217_ better discriminates amyloid-positive individuals, partly explaining the observed performance gap^42^.

**Figure 6.**
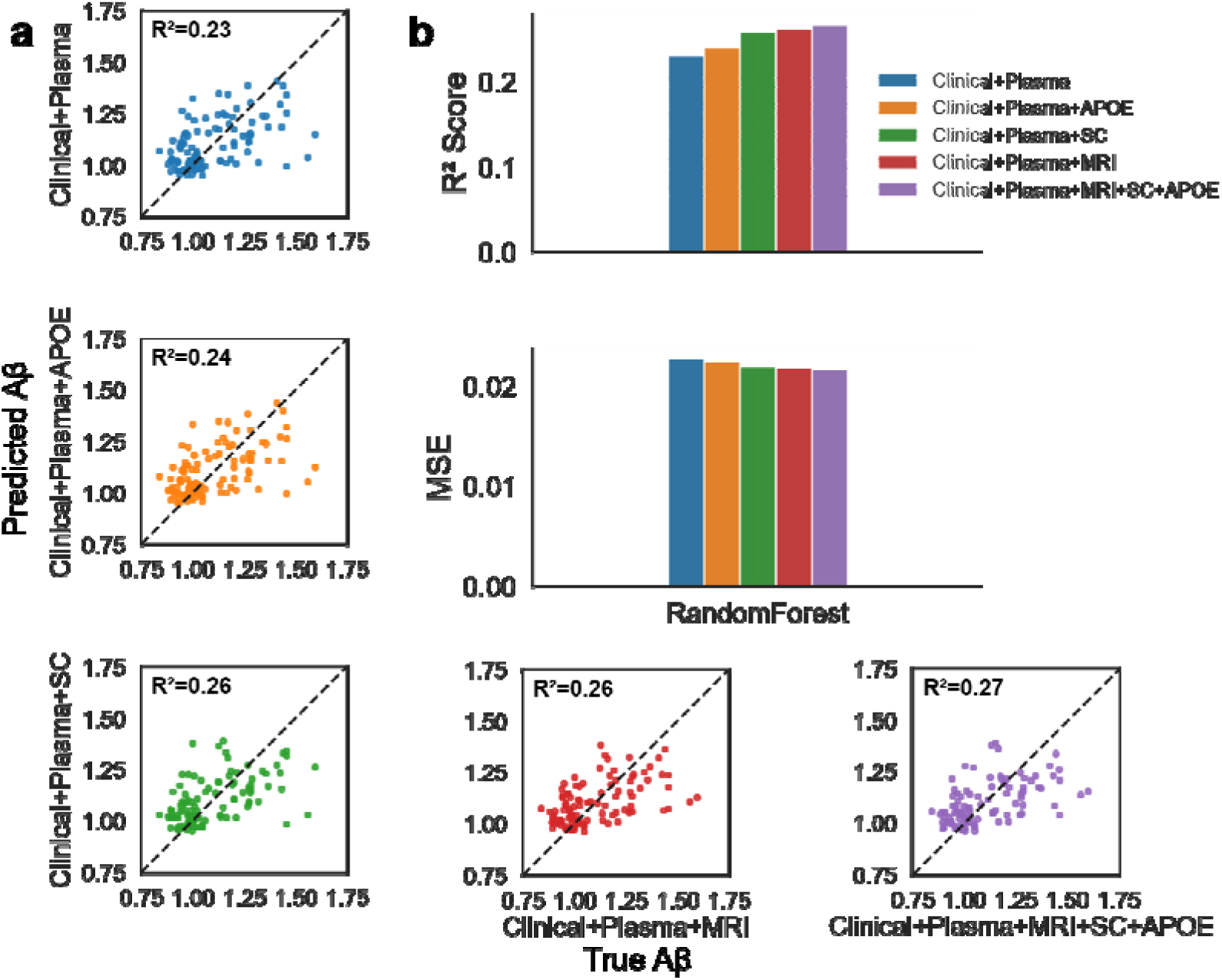
Model performance for Aβ burden prediction in the SILCODE cohort. (a) Scatter plots of observed vs. predicted Aβ SUVR values across five feature combinations using RF models. R² values indicate model fit. The x-axis denotes observed SUVR values; the y-axis shows model-predicted values. (b) Bar plots summarize model performance under a LOOCV framework. The upper panel shows R² values, and the lower panel displays MSE across feature combinations. Abbreviations: Aβ, amyloid-β; APOE, apolipoprotein E; GFAP, glial fibrillary acidic protein; MMSE, Mini-Mental State Examination; NfL, neurofilament light chain; PCA, principal component analysis; SC, structural connectivity; RF, random forest; MSE, mean squared error. Clinical, demographic and cognitive measures.

### 3.6 Sensitivity Analysis

First, we tested multiple p-value thresholds (p < 10^-5^ to 10^-10^) for PRS construction using the clumping-and-thresholding approach. All thresholds except p < 10^-5^ improved prediction over the plasma-only baseline, with the best performance observed at p < 10^-6^. These results underscore the importance of optimal thresholding for leveraging genome-wide polygenic data (Table S5 and Fig S1). Second, we evaluated the impact of different parcellation schemes on SC-based prediction. Alternative atlases with 84 and 116 regions both outperformed the baseline, with minor differences(Table S6 and Fig S2). These findings suggest that SC features provide stable predictive value across anatomical resolutions, with higher-resolution schemes offering more refined disease-relevant connectivity patterns. Together, these analyses show that predictive gains from integrating imaging and genetic features are robust to key modeling and design choices.

## 4 Discussion

We proposed a multimodal machine learning framework that integrates plasma biomarkers, sMRI, DTI-derived structural connectivity (SC), and genetic risk to non-invasively predict cerebral Aβ burden. In the ADNI cohort, adding sMRI, SC features, and APOE genotype to clinical and plasma measures substantially improved model performance (R² = 0.617 vs. R² = 0.515 for clinical + plasma only). Replacing APOE with a genome-wide polygenic risk score (PRS) further enhanced predictive accuracy (R² = 0.637), highlighting the added value of capturing distributed genetic risk beyond the APOE locus^10,35^. These results underscore the complementary strengths of different modalities in characterizing early Alzheimer’s disease pathology. External validation in the SILCODE cohort supported the generalizability of our findings. Despite differences in demographics, imaging protocols, and biomarker assays, consistent multimodal predictive improvements across cohorts underscore the framework’s generalizability and translational potential.

Beyond continuous prediction, our multimodal model also showed improved classification of individuals along the AD spectrum. Compared to the clinical + plasma model, the full model achieved higher AUC scores across all diagnostic stages (NC: 0.86; MCI: 0.77; AD: 0.93), reinforcing its utility for both quantitative and categorical assessments of Aβ burden.

Structurally, sMRI and SC features offered complementary insights into disease mechanisms. While sMRI captured diffuse cortical atrophy, SC analysis revealed spatially organized disconnection patterns. Notably, the visual network exhibited the highest number of disrupted connections, both internally and with the control, somatomotor, and default mode networks—suggesting early vulnerability of perceptual and integrative attention systems in preclinical AD^25^. In addition, disconnections frequently involved the medial prefrontal and lateral parietal cortices—core hubs of the default mode and frontoparietal networks—coinciding with known regions of early Aβ deposition^43^. These findings support a network-level interpretation of amyloid pathology and underscore the utility of CPM-derived SC features as sensitive and interpretable markers of early brain changes.

From a genetic perspective, APOE ε4 consistently demonstrated predictive utility across cohorts. However, PRS outperformed APOE in the ADNI dataset, likely due to its capacity to aggregate genome-wide susceptibility variants^10^. Despite its superior predictive power, PRS remains less accessible and more costly, making APOE genotyping a more pragmatic option in resource-limited clinical settings^8^.

Sensitivity analyses confirmed that our findings were robust across different PRS p-value thresholds and SC parcellation schemes. Nevertheless, several limitations warrant consideration. First, our sample sizes were modest, which may restrict the generalizability of the model to broader populations^44^. Second, the use of the Schaefer-100/MSA-16 atlas prioritized interpretability but may have limited our ability to detect finer-grained network effects^25^. Future work should explore higher-resolution or individualized parcellations to refine SC modeling.

Implications. Collectively, our study demonstrates that integrating multimodal information—including plasma biomarkers, neuroimaging, and genetic risk—can significantly enhance non-invasive prediction of cerebral amyloid pathology. This scalable framework holds potential for improving early AD risk stratification and may inform future screening strategies in both clinical and population-based settings.

## Ethics declarations

All procedures performed in studies involving human participants were in accordance with the ethical standards of the institutional and/or national research committees and with the 1964 Declaration of Helsinki and its later amendments or comparable ethical standards.

For the ADNI cohort, ethical approval was obtained from the institutional review boards of all participating institutions (e.g., University of California, San Diego). For the SILCODE cohort, the study was approved by the Ethics Committee of Xuanwu Hospital, Capital Medical University. Written informed consent was obtained from all participants in both cohorts.

## Consent for publication

Not applicable. No individual person’s data is included in this study.

## Data availability

The data used in this study are publicly available from the Alzheimer’s Disease Neuroimaging Initiative (ADNI; http://adni.loni.usc.edu) and the SILCODE data are available upon reasonable request from the corresponding author.

## Acknowledgments

We thank all study participants and their families for their generous contributions. We are grateful to the Alzheimer’s Disease Neuroimaging Initiative (ADNI) investigators and staff for participant recruitment and data acquisition, and to the SILCO DE research team for providing external□validation data.

## References

1. Selkoe, D. J. & Hardy, J. The amyloid hypothesis of Alzheimer’s disease at 25 years. EMBO Mol Med 8, 595–608 (2016).

2. DeTure, M. A. & Dickson, D. W. The neuropathological diagnosis of Alzheimer’s disease. Molecular Neurodegeneration 14, 32 (2019).

3. Jack, C. R. et al. NIA-AA Research Framework: Toward a biological definition of Alzheimer’s disease. Alzheimers Dement 14, 535–562 (2018).

4. Nakamura, A. et al. High performance plasma amyloid-β biomarkers for Alzheimer’s disease. Nature 554, 249–254 (2018).

5. Zhang, M. et al. Relationship between topological efficiency of white matter structural connectome and plasma biomarkers across the Alzheimer’s disease continuum. Human Brain Mapping 45, e26566 (2024).

6. Racine, A. M. et al. Associations between white matter microstructure and amyloid burden in preclinical Alzheimer’s disease: A multimodal imaging investigation. NeuroImage: Clinical 4, 604–614 (2014).

7. Huang, W. et al. Individual Variability in the Structural Connectivity Architecture of the Human Brain. J Neurosci 45, e2139232024 (2025).

8. Liu, C.-C., Liu, C.-C., Kanekiyo, T., Xu, H. & Bu, G. Apolipoprotein E and Alzheimer disease: risk, mechanisms and therapy. Nat Rev Neurol 9, 106–118 (2013).

9. Leonenko, G. et al. Identifying individuals with high risk of Alzheimer’s disease using polygenic risk scores. Nat Commun 12, 4506 (2021).

10. Ramanan, V. K. et al. Genetic risk scores enhance the diagnostic value of plasma biomarkers of brain amyloidosis. Brain 146, 4508–4519 (2023).

11. Karlsson, L. et al. Machine learning prediction of taulPET in Alzheimer’s disease using plasma, MRI, and clinical data.

12. Ying, Q. et al. Multi-Modal Data Analysis for Alzheimer’s Disease Diagnosis: An Ensemble Model Using Imagery and Genetic Features. Annu Int Conf IEEE Eng Med Biol Soc 2021, 3586–3591 (2021).

13. Sarica, A., Cerasa, A. & Quattrone, A. Random Forest Algorithm for the Classification of Neuroimaging Data in Alzheimer’s Disease: A Systematic Review. Front Aging Neurosci 9, 329 (2017).

14. Santos, L. E. et al. Performance of plasma biomarkers for diagnosis and prediction of dementia in a Brazilian cohort. Nat Commun 16, 2911 (2025).

15. Identifying incipient dementia individuals using machine learning and amyloid imaging. Neurobiology of Aging 59, 80–90 (2017).

16. Pais, M. V., Forlenza, O. V. & Diniz, B. S. Plasma Biomarkers of Alzheimer’s Disease: A Review of Available Assays, Recent Developments, and Implications for Clinical Practice. J Alzheimers Dis Rep 7, 355–380.

17. Alfaro-Almagro, F. et al. Image processing and Quality Control for the first 10,000 brain imaging datasets from UK Biobank. NeuroImage 166, 400–424 (2018).

18. Jenkinson, M., Beckmann, C. F., Behrens, T. E., Woolrich, M. W. & Smith, S. M. FSL. NeuroImage 62, 782–790 (2012).

19. Fischl, B. FreeSurfer. NeuroImage 62, 774–781 (2012).

20. Tournier, J.-D. et al. MRtrix3: A fast, flexible and open software framework for medical image processing and visualization. NeuroImage 202, 116137 (2019).

21. Li, X., Morgan, P. S., Ashburner, J., Smith, J. & Rorden, C. The first step for neuroimaging data analysis: DICOM to NIfTI conversion. Journal of Neuroscience Methods 264, 47–56 (2016).

22. Andersson, J. L. R. & Sotiropoulos, S. N. An integrated approach to correction for off-resonance effects and subject movement in diffusion MR imaging. Neuroimage 125, 1063–1078 (2016).

23. Tournier, J.-D., Calamante, F. & Connelly, A. Robust determination of the fibre orientation distribution in diffusion MRI: Non-negativity constrained super-resolved spherical deconvolution. NeuroImage 35, 1459–1472 (2007).

24. Smith, R. E., Tournier, J.-D., Calamante, F. & Connelly, A. Anatomically-constrained tractography: Improved diffusion MRI streamlines tractography through effective use of anatomical information. NeuroImage 62, 1924–1938 (2012).

25. Schaefer, A. et al. Local-global parcellation of the human cerebral cortex from intrinsic functional connectivity MRI. Cerebral Cortex 28, 3095–3114 (2018).

26. Tian, Y., Margulies, D. S., Breakspear, M. & Zalesky, A. Hierarchical organization of the human subcortex unveiled with functional connectivity gradients. Nature Neuroscience 23, 1421–1432 (2020).

27. Landau, S. M. et al. Amyloid deposition, hypometabolism, and longitudinal cognitive decline. Annals of Neurology 72, 578–586 (2012).

28. Klunk, W. E. et al. The Centiloid Project: standardizing quantitative amyloid plaque estimation by PET. Alzheimers Dement 11, 1–15.e1–4 (2015).

29. Chen, H.-J. et al. Amyloid pathology related to aberrant structure-function coupling of brain networks in Alzheimer’s disease: insights from [18F]-florbetapir PET imaging. Eur J Nucl Med Mol Imaging (2025) doi:10.1007/s00259-025-07172-8.

30. Clark, C. M. et al. Use of florbetapir-PET for imaging beta-amyloid pathology. JAMA 305, 275–283 (2011).

31. Purcell, S. et al. PLINK: a tool set for whole-genome association and population-based linkage analyses. Am J Hum Genet 81, 559–575 (2007).

32. Danecek, P. et al. Twelve years of SAMtools and BCFtools. Gigascience 10, giab008 (2021).

33. Auton, A. et al. A global reference for human genetic variation. Nature 526, 68–74 (2015).

34. Rubinacci, S., Ribeiro, D. M., Hofmeister, R. J. & Delaneau, O. Efficient phasing and imputation of low-coverage sequencing data using large reference panels. Nat Genet 53, 120–126 (2021).

35. Jansen, I. E. et al. Genome-wide meta-analysis identifies new loci and functional pathways influencing Alzheimer’s disease risk. Nat Genet 51, 404–413 (2019).

36. Prakash, R. S. et al. A whole-brain functional connectivity model of Alzheimer’s disease pathology. Alzheimers Dement 21, e14349 (2025).

37. Fjell, A. M. et al. One-year brain atrophy evident in healthy aging. J Neurosci 29, 15223–15231 (2009).

38. Finn, E. S. Functional connectome fingerprinting: identifying individuals using patterns of brain connectivity. nature NEUROSCIENCE 18, (2015).

39. Shen, X. et al. Using connectome-based predictive modeling to predict individual behavior from brain connectivity. Nat Protoc 12, 506–518 (2017).

40. Varoquaux, G. Cross-validation failure: Small sample sizes lead to large error bars. Neuroimage 180, 68–77 (2018).

41. Lundberg, S. M. & Lee, S.-I. A unified approach to interpreting model predictions. In Advances in Neural Information Processing Systems vol. 30 4765–4774 (2017).

42. Palmqvist, S. et al. Discriminative Accuracy of Plasma Phospho-tau217 for Alzheimer Disease vs Other Neurodegenerative Disorders. JAMA 324, 772–781 (2020).

43. Thal, D. R., Rüb, U., Orantes, M. & Braak, H. Phases of Aβ-deposition in the human brain and its relevance for the development of AD. Neurology 58, 1791–1800 (2002).

44. Luckett, E. S. et al. Association of Alzheimer’s disease polygenic risk scores with amyloid accumulation in cognitively intact older adults. Alzheimers Res Ther 14, 138 (2022).

