## Supplemental Data 1 for "Multimodal Integration of Alzheimer’s Plasma Biomarkers, MRI, and Genetic Risk for Individual Prediction of Cerebral Amyloid Burden"

| **Frontal regions:** | **Anterior/posterior cingulate regions:** | **Lateral parietal regions:** | **Lateral temporal regions:** |
| --- | --- | --- | --- |
| ctx-lh-caudalmiddlefrontal | ctx-lh-caudalanteriorcingulate | ctx-lh-inferiorparietal | ctx-lh-inferiortemporal |
| ctx-lh-lateralorbitofrontal | ctx-lh-isthmuscingulate | ctx-lh-precuneus | ctx-lh-middletemporal |
| ctx-lh-medialorbitofrontal | ctx-lh-posteriorcingulate | ctx-lh-superiorparietal | ctx-lh-superiortemporal |
| ctx-lh-parsopercularis | ctx-lh-rostralanteriorcingulate | ctx-lh-supramarginal | ctx-rh-inferiortemporal |
| ctx-lh-parsorbitalis | ctx-rh-caudalanteriorcingulate | ctx-rh-inferiorparietal | ctx-rh-middletemporal |
| ctx-lh-parstriangularis | ctx-rh-isthmuscingulate | ctx-rh-precuneus | ctx-rh-superiortemporal |
| ctx-lh-rostralmiddlefrontal | ctx-rh-posteriorcingulate | ctx-rh-superiorparietal |  |
| ctx-lh-superiorfrontal | ctx-rh-rostralanteriorcingulate | ctx-rh-supramarginal |  |
| ctx-lh-frontalpole |  |  |  |
| ctx-rh-caudalmiddlefrontal |  |  |  |
| ctx-rh-lateralorbitofrontal |  |  |  |
| ctx-rh-medialorbitofrontal |  |  |  |
| ctx-rh-parsopercularis |  |  |  |
| ctx-rh-parsorbitalis |  |  |  |
| ctx-rh-parstriangularis |  |  |  |
| ctx-rh-rostralmiddlefrontal |  |  |  |
| ctx-rh-superiorfrontal |  |  |  |
| ctx-rh-frontalpole |  |  |  |

Supplementary Table 1. Cortical regions used to compute the summary Aβ-PET SUVR.

Regions were defined according to the Desikan–Killiany atlas and grouped into four macroscale anatomical categories: frontal cortex, anterior/posterior cingulate cortex, lateral parietal cortex, and lateral temporal cortex. The summary SUVR was computed as the mean tracer uptake across these regions, following co-registration of native-space PET to FreeSurfer-segmented MRI, and normalized to the whole cerebellum. Region names follow FreeSurfer conventions.

R2

|  | Clinical+Plasma | Clinical+Plasma+APOE | Clinical+Plasma+SC | Clinical+Plasma+sMRI | Clinical+Plasma+sMRI+SC+APOE |
| --- | --- | --- | --- | --- | --- |
| Linear | 0.358559 | 0.394536 | 0.363004 | 0.342823 | 0.380436 |
| RandomForest | 0.515303 | 0.527164 | 0.556337 | 0.614673 | 0.616606 |

mse

|  | Clinical+Plasma | Clinical+Plasma+APOE | Clinical+Plasma+SC | Clinical+Plasma+sMRI | Clinical+Plasma+sMRI+SC+APOE |
| --- | --- | --- | --- | --- | --- |
| Linear | 1301.016 | 1228.044 | 1292 | 1332.933 | 1256.643 |
| RandomForest | 983.0964 | 959.0393 | 899.8689 | 781.5475 | 777.6271 |

Supplementary Table 2. Prediction performance of different feature combinations for Aβ burden (Centiloid) estimation.

The table reports the coefficient of determination (R²) and mean squared error (MSE) for linear regression and random forest (RF) models under a leave-one-out cross-validation (LOOCV) framework. Five feature sets were evaluated: (1) Clinical + Plasma, (2) Clinical + Plasma + APOE, (3) Clinical + Plasma + Structural Connectivity (SC), (4) Clinical + Plasma + Structural MRI (sMRI), and (5) Clinical + Plasma + sMRI + SC + APOE. While linear regression showed limited predictive power across all combinations, RF models achieved progressively better performance with the inclusion of multimodal features. The best model integrated all modalities, yielding an R² of 0.617 and the lowest MSE (≈777.6).

R2

|  | Clinical+Plasma | Clinical+Plasma+APOE | Clinical+Plasma+PRS | Clinical+Plasma+MRI+SC+APOE | Clinical+Plasma+MRI+SC+PRS |
| --- | --- | --- | --- | --- | --- |
| RandomForest | 0.515303 | 0.527164 | 0.571467 | 0.616727 | 0.637398 |

MSE

|  | Clinical+Plasma | Clinical+Plasma+APOE | Clinical+Plasma+PRS | Clinical+Plasma+MRI+SC+APOE | Clinical+Plasma+MRI+SC+PRS |
| --- | --- | --- | --- | --- | --- |
| RandomForest | 983.0964 | 959.0393 | 869.1816 | 777.3805 | 735.4542 |

Supplementary Table 3. Comparative performance of APOE genotype and polygenic risk score (PRS 1 × 10^-6^) in predicting Aβ burden (Centiloid).

This table presents the R² and mean squared error (MSE) of random forest models trained on different combinations of clinical, plasma, neuroimaging, and genetic features for predicting cortical Centiloid values in the ADNI cohort. The inclusion of APOE genotype in the baseline Clinical + Plasma model yielded a modest performance gain (R² = 0.527; MSE = 959.0). In contrast, replacing APOE with polygenic risk scores (PRS) resulted in a larger improvement (R² = 0.571; MSE = 869.2), highlighting the additive predictive value of genome-wide genetic information beyond APOE alone. Models incorporating both neuroimaging features (sMRI + SC) and genetic variables showed further enhancement, with the full model containing PRS achieving the highest predictive performance (R² = 0.637; MSE = 735.5).

R2

|  | Clinical+Plasma | Clinical+Plasma+APOE | Clinical+Plasma+SC | Clinical+Plasma+MRI | Clinical+Plasma+MRI+SC+APOE |
| --- | --- | --- | --- | --- | --- |
| RandomForest | 0.231584 | 0.240094 | 0.259256 | 0.263085 | 0.267136 |

MSE

|  | Clinical+Plasma | Clinical+Plasma+APOE | Clinical+Plasma+SC | Clinical+Plasma+MRI | Clinical+Plasma+MRI+SC+APOE |
| --- | --- | --- | --- | --- | --- |
| RandomForest | 0.022718 | 0.022466 | 0.0219 | 0.021786 | 0.021667 |

Supplementary Table 4. External validation of Aβ burden prediction in the SILCODE cohort.

This table summarizes the predictive performance (R² and mean squared error, MSE) of random forest models applied to the independent SILCODE dataset under five multimodal feature combinations. All models aimed to predict regional Aβ-PET SUVR values. Starting from the baseline model (Clinical + Plasma), the incremental inclusion of APOE genotype, structural connectivity (SC), and structural MRI (sMRI) features yielded consistent performance improvements. The full model incorporating all modalities achieved the highest R² (0.267) and the lowest MSE (≈0.0217), highlighting the generalizability of the proposed framework.

R2

|  | Clinical+Plasma | Clinical+Plasma+PRS_1e-05 | Clinical+Plasma+PRS_1e-06 | Clinical+Plasma+PRS_1e-07 | Clinical+Plasma+PRS_1e-08 | Clinical+Plasma+PRS_1e-09 | Clinical+Plasma+PRS_1e-10 |
| --- | --- | --- | --- | --- | --- | --- | --- |
| RandomForest | 0.515303 | 0.511209 | 0.571467 | 0.536659 | 0.540494 | 0.553087 | 0.560889 |

MSE

|  | Clinical+Plasma | Clinical+Plasma+PRS_1e-05 | Clinical+Plasma+PRS_1e-06 | Clinical+Plasma+PRS_1e-07 | Clinical+Plasma+PRS_1e-08 | Clinical+Plasma+PRS_1e-09 | Clinical+Plasma+PRS_1e-10 |
| --- | --- | --- | --- | --- | --- | --- | --- |
| RandomForest | 983.0964 | 991.4012 | 869.1816 | 939.7805 | 932.0036 | 906.4605 | 890.6367 |

Supplementary Table 5. Impact of p-value thresholds for PRS construction on Aβ burden (Centiloid) prediction.

Prediction performance (R² and MSE) of random forest models using Clinical + Plasma + PRS features is shown across a range of PRS p-value thresholds (p < 10⁻⁵ to 10⁻¹⁰), based on the clumping-and-thresholding approach. The bar plots and table demonstrate that all thresholds except p < 10⁻⁵ improved prediction over the Clinical + Plasma baseline, with the best performance achieved at p < 10⁻⁶ (R² = 0.571; MSE = 869.2). These findings highlight the importance of optimal thresholding for maximizing the predictive power of genome-wide polygenic scores in estimating cerebral Aβ burden.

R2

|  | Clinical+Plasma | Clinical+Plasma+SC84 | Clinical+Plasma+SC116 |
| --- | --- | --- | --- |
| RandomForest | 0.515303 | 0.556337 | 0.56675 |

MSE

|  | Clinical+Plasma | Clinical+Plasma+SC84 | Clinical+Plasma+SC116 |
| --- | --- | --- | --- |
| RandomForest | 983.0964 | 899.8689 | 878.7494 |

Supplementary Table 6. Influence of parcellation schemes on SC-based prediction of Aβ burden (Centiloid).

To evaluate the robustness of structural connectivity (SC) features across anatomical resolutions, random forest models were constructed using SC matrices generated from two parcellation schemes: an 84-region and a 116-region atlas. Both feature sets were combined with clinical and plasma biomarkers to predict Centiloid values. As shown in the figure and table, both SC84 and SC116 configurations outperformed the Clinical + Plasma baseline (R² = 0.556 and 0.567 vs. 0.515), with modest improvements in predictive accuracy and reductions in MSE. These results suggest that SC-based features contribute stably across parcellation schemes and that higher-resolution atlases may offer slight advantages in delineating disease-relevant connectivity patterns.
