## Supplemental Data 2 for "Multimodal Integration of Alzheimer’s Plasma Biomarkers, MRI, and Genetic Risk for Individual Prediction of Cerebral Amyloid Burden"

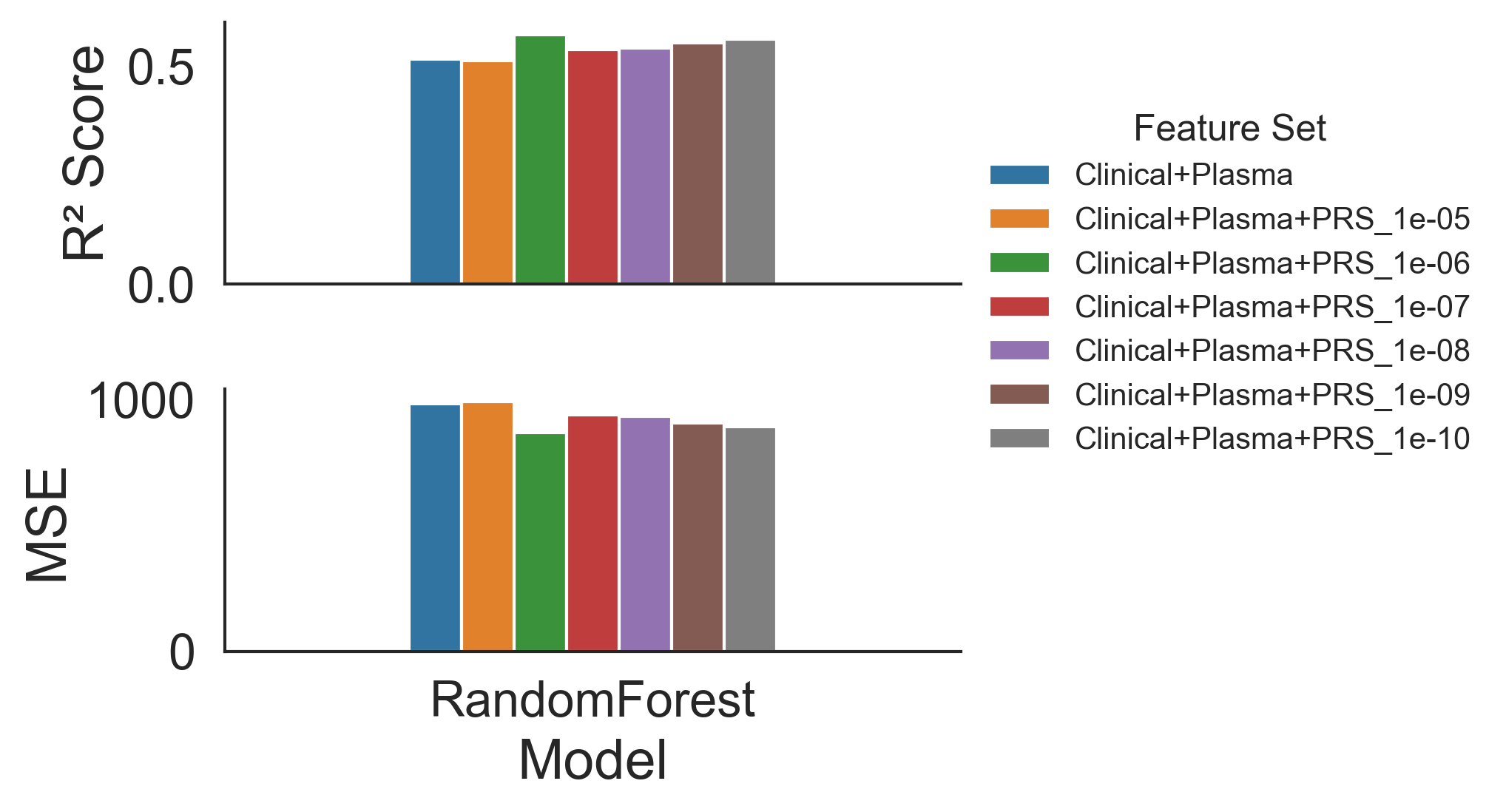

Supplementary Fig 1. Impact of p-value thresholds for PRS construction on Aβ burden (Centiloid) prediction.

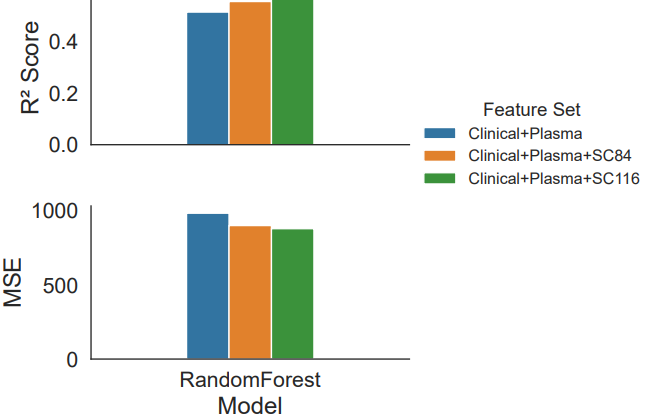

Supplementary Fig 2. Influence of parcellation schemes on SC-based prediction of Aβ burden (Centiloid).
